# Evolutionary and transmission dynamics of vaccine-derived poliovirus serotype 2 (cVDPV2) outbreaks in the Horn of Africa region

**DOI:** 10.64898/2026.11.25.26368672

**Authors:** D. Collins Owuor, Collins K. Cheruiyot, Mercy Onyango, Stephen O. Ombija, Shadrack Barmasai, Rosemary Nzunza, Joanne Hassan, Samira Ali, Agnes Chepkurui, Agnes Ndunge, Evans Komen, Benlick Mwangi, Lawrence O. Oswago, Cheroigin Kipchirchir, Carolyne Gathenji, John Kipterer, Michael N. Mwanza, Kebba Touray, Asma Ali, Rennatus Mdodo, Samoel A. Khamadi, Peter Borus

## Abstract

**Background:** The poliovirus eradication campaign by the Global Polio Eradication Initiative (GPEI) has resulted in the eradication of wild poliovirus (WPV) serotypes two (WPV2) and three (WPV3) and limited transmission of serotype one (WPV1) in two countries. However, increased emergence of circulating vaccine-derived poliovirus (cVDPV) outbreaks and continued circulation of WPV1 poses a continuous threat of international spread. Recently, cVDPV serotype 2 (cVDPV2) isolates from environmental samples in five World Health Organization (WHO) European Region (EURO) countries were genetically linked to cVDPV2 originating in Nigeria.

**Methods:** We utilized phylogeographic methods to infer the origins and geographic spread cVDPV2 poliovirus in WHO African Region (AFRO) Horn of Africa using 234 full-length VP1 gene sequences from three WHO AFRO and one WHO Eastern Mediterranean Region (EMRO) countries collected through clinical and environmental surveillance between 2017 and 2025.

**Findings:** cVDPV2 outbreak strains from different countries in the Horn of Africa spanned the overall SOM-BAN-1 phylogeny, reflecting multiple introductions into the region, with strains harboring unique nucleotide mutations and amino acid substitutions. Phylogeographic analyses revealed SOM-BAN-1 origins from Somalia, with subsequent spread to other Horn of African countries and subsequent seeding of cVDPV2 outbreaks from other countries in the region, but with phylogenetic predominance of Somalia. The cVDPV2 transmission dynamics in the region were characterized by circulation of multiple chains of transmission in multiple countries, with poliovirus spread characterized by localized evolution and regionally linked chains of transmission. Stronger evidence for SOM-BAN-1 origins from Somalia was observed, with multiple chains of transmission observed in multiple sub-national locations, supporting the important role played by the country in the predominance and persistence of cVDPV2 in Horn of Africa region.

**Interpretation:** The spread of cVDPV2 outside the country of first detection, with further international spread, reveals gaps in the effectiveness and timeliness of outbreak responses. Therefore, regular updates of poliovirus surveillance sequence data using phylogeographic tools may provide insights into changes in poliovirus epidemiology, which can in turn guide the implementation of timely outbreak response and more strategic and effective vaccination campaigns. The analyses may be particularly relevant to understand the evolutionary and transmission dynamics of ongoing cVDPV outbreaks in AFRO, halting ongoing transmissions in AFRO and recent spread to other regions, achieving the GPEIs efforts of a poliovirus-free world.

## Introduction

The emergence and spread of vaccine-derived polioviruses (VDPVs), which are oral polio vaccine (OPV) strains (or Sabin strains) that have undergone prolonged circulation in under-vaccinated populations resulting in genetic reversion to neurovirulence, is a major hurdle towards a poliovirus-free world [1]. Outbreaks of VDPVs resulted in a global withdrawal of serotype two OPV through a globally coordinated switch from trivalent (tOPV) to bivalent OPV (bOPV) in April 2016 [2, 3]. However, challenges with bOPV vaccination coverage have resulted in a growing cohort of children unimmunized against serotype two poliovirus particularly in the African Region (AFRO). In summary, there was an increasing incidence of serotype two VDPV (VDPV2) outbreaks, emergent from trivalent OPV (tOPV) vaccines used before the switch and monovalent serotype two OPV (mOPV2) vaccines used to control VDPV2 outbreaks [4, 5]. A novel type 2 OPV (nOPV2) was developed, with low probability of reversion to paralytic variants than mOPV2/tOPV2 and limited genetic modifications when used [6]. However, reversions in nOPV2 have been reported, but less frequent compared to mOPV2 and tOPV2 [7].

While wild poliovirus (WPV) is the most known form of poliovirus, circulating VDPV (cVDPV) is a form that can spread within communities with low immunization coverage. Although rare, cVDPV outbreaks have been increasing in recent years due to low immunization rates within communities, with cVDPV type 2 (cVDPV2) outbreaks being the most prevalent [6], particularly in World Health Organization (WHO) AFRO [8–10]. With WHO AFRO declared to have interrupted transmission of WPV in August 2020, cVDPV outbreaks are now the only form of the poliovirus that affects the region, as new outbreaks continue to be reported in multiple countries [6, 10]. For example, during January 2023 – June 2024, a total of 74 cVDPV2 outbreaks from 34 emergences were reported in 38 countries, 31 of which were countries in WHO AFRO [9]. During the reporting period, cVDPV outbreaks (≥12 months from first to most recent detection) were observed in 15 countries, 12 of which were countries in AFRO [9]. However, a WPV1 virus was isolated from a stool sample collected in Malawi in November 2021 and in Mozambique in March 2022 [11]. Additionally, cVDPV2 isolates from environmental samples in five WHO European Region (EURO) countries (Finland, Germany, Poland, Spain, and the United Kingdom) were recently genetically linked to cVDPV2 strains originating in Zamfara, Nigeria [12]. The importation of WPV1 into WHO AFRO and recent exportation of cVDPV2 from WHO AFRO to WHO EURO highlights the continued threat posed by poliovirus globally and the need to further strengthen surveillance and outbreak response particularly in AFRO. Additionally, there is need for novel strategies to guide implementation of more strategic and effective vaccination campaigns.

Improved understanding of the origins and geographic spread of cVDPVs, through analysis of sequence data provides a better understanding of virus transmission, which can in turn guide the implementation of timely outbreak response and more strategic and effective vaccination campaigns [8, 13]. Genetic sequencing of cVDPVs detected through clinical and environmental surveillance is useful for confirmation of virus detection, identification of cVDPV origins, tracking geographic patterns of spread, and determination of appropriate vaccination responses. For example, it is possible to infer which of cVDPV outbreak cases are due to local transmissions and which are more likely to be linked to chains of cVDPV transmission in other countries. A recent study linked cVDPV2 outbreak cases in Benin, Cameroon, Ghana, and Niger to ongoing circulation in Northern Nigeria, whereas cases in Angola and Democratic Republic of the Congo resulted from unlinked local emergences [14]. Understanding which cases are due to under-vaccinated local populations and which are due to international spread is important in the determination of a correct vaccine response and to investigate the cause of the outbreak [13].

This current study utilized phylodynamics methods based on Nextstrain real-time tracking of pathogen evolution [15, 16] and Delphy Bayesian phylogenetics [17] to infer the origins and geographic spread of cVDPVs between and within AFRO countries in the Horn of Africa. Regular updates of the cVDPV surveillance sequence data using phylogeographic visualization tools may provide insights into changes in cVDPV epidemiology, which can in turn guide the implementation of more strategic and effective Supplementary Immunization Activities (SIA). These analyses may be particularly relevant to interrupt the ongoing outbreaks of cVDPVs in WHO AFRO through improved surveillance and timely and effective outbreak response, achieving the Global Polio Eradication Initiative’s (GPEIs) efforts of a poliovirus-free world.

## Materials and Methods

### Study setting and data sources

The study was conducted in WHO AFRO, one of the six WHO regions that includes 47 countries in the African continent (**Figure S1A**). The region includes diverse geographical areas with diverse cultural backgrounds and socioeconomic conditions. The main data source was the WHO Polio Information System (POLIS), managed centrally at the WHO headquarters, a comprehensive platform that tracks poliovirus related indicators including confirmed cases, surveillance indicators, laboratory sequencing results and SIAs [18]. Data were extracted from POLIS on July 17, 2026, in Comma-Separated Values (CSV) file format. The dataset included multiple variables such as demographics, sample source (acute flaccid paralysis (AFP), environmental, contact), laboratory test results, and genetic sequence data. Where possible, socioeconomic and clinical information were triangulated with laboratory metadata to give a comprehensive profile of each sampled virus.

### Poliovirus surveillance in WHO AFRO

Poliovirus surveillance is one of the GPEI’s strategies for eradication of the virus. As the GPEI comes closer to the global goal of WPV eradication, sensitive surveillance allowing to reliably conclude the absence of poliovirus circulation becomes increasingly important. To date, poliovirus surveillance permitting the reliable and timely detection of all types of polioviruses (WPV, VDPV, Sabin-like viruses) is mainly conducted using AFP and environmental surveillance [19]. Recently this was complemented by a third approach referred to as immunodeficiency-associated vaccine-derived poliovirus (iVDPV) surveillance among primary immunodeficient subjects. AFP surveillance is the gold standard for detecting polio cases. The goal of AFP surveillance is to identify and document the presence or absence of poliovirus. AFP surveillance is the process of detecting, reporting and investigating AFP cases in children below 15 years of age or in any person of any age when a clinician suspects poliomyelitis, while environmental surveillance is the routine collection and testing of sewage samples for poliovirus in specific settings [20].

### Sample collection, laboratory and sequencing procedures

Two stool samples, collected 24–48 hours apart from AFP cases and sewage samples treated according to the standard protocols, were cultured on L20B cells (mouse L cells expressing the human CD155 poliovirus receptor) and RD cells (human rhabdomyosarcoma cells) for viral detection, following WHO guidelines [21, 22]. Poliovirus was suspected when isolates were detected in both RD and L20B cell lines. These suspected poliovirus isolates were further analyzed through intratypic differentiation (ITD) using real-time reverse transcriptase PCR (rRT-PCR) and a poliovirus diagnostic kit to identify serotypes and distinguish wild polioviruses, in alignment with the GPEI framework [23]. Sabin-related polioviruses were subsequently screened for VDPVs using rRT-PCR [23, 24]. All non-Sabin-like poliovirus isolates underwent genomic sequencing for further characterization [24–26].

### Spatiotemporal analyses of poliovirus sequence datasets

Spatiotemporal analyses were conducted to characterize the spatial and temporal distribution of cVDPV2 SOM-BAN-1 emergence group polioviruses, which were the most predominant and persistent emergence group in WHO AFRO Horn of Africa region during the study period, 2017-2025. SOM-BAN-1 was first detected in Somalia in 2017 [9], spreading to 4 other countries in the Horn of Africa following emergence as reported in the WHO AFRO Polio eradication program in the polio detections database (https://afrodim.shinyapps.io/new-virus-detections/); the emergence groups was last reported in October 2025. The emergence group was selected to characterize the evolutionary and transmission dynamics of cVDPV2 in the region. The spatiotemporal analysis was characterized and visualized in R Studio (https://posit.co/downloads/).

Temporal changes in the relative frequency of cVDPV2 sequences by Horn of Africa country were assessed using a country-stratified frequency analysis over the study period. Sequence data were grouped by country of origin and sampling year, with the proportion of sequences contributed by each country calculated for each time point. The resulting frequencies were expressed as percentages of total sequences sampled within each time interval and visualized using a stacked area plot. This approach enables comparison of the temporal distribution and relative contribution of Somalia, Kenya, and Ethiopia to the overall sequence dataset.

### Phylogenetic reconstruction

Full-length VP1 gene sequences (903 base pairs) of cVDPV2 from four WHO AFRO countries in the Horn of Africa, collected between 2017 and 2025, were utilized for the phylogenetic analysis. Raw and time-scaled phylogenomic trees were built using the Nextstrain pipeline (https://github.com/nextstrain, command-line interface (CLI) version 11.1.2) [15, 16] installed in a Conda environment (https://github.com/conda/conda, version 26.7.0) [16]. The pipeline utilizes the Augur toolkit (https://github.com/nextstrain/augur) for alignment, phylogenetic inference, temporal dating of ancestral nodes, ancestral location state reconstruction (based on discrete phylogeographic reconstruction), and inference of the most likely transmission events. The pipeline generates an output file processed by the Auspice web interface (https://github.com/nextstrain/auspice, version 2.73.0) [15, 16] to explore phylodynamic and phylogenomic data. The Augur/align module was deployed to execute the multiple sequence alignment with MAFFT (https://github.com/GSLBiotech/mafft, version v7.526) [27] using poliovirus serotype 2 (Sabin 2) as a reference genome. We then used IQ-TREE (http://www.iqtree.org/, version 3.1.2) [28] to reconstruct a phylogenetic tree of VP1 sequences from the WHO AFRO Horn of Africa and Eastern Mediterranean Region (EMRO) countries, based on the general time reversible (GTR) model of nucleotide substitution and a proportion of gamma (G) and invariable sites (+I), with bootstrap method of 1000 replicates. The final phylogenetic inferrence was conducted following the removal of outlier strains, as characterized by TempEst (http://tree.bio.ed.ac.uk/software/tempest/) to evaluate the temporal phylogenetic signal in the data, based on root-to-tip divergence estimates.

### Phylogeographic reconstruction

Branch lengths, sampling dates, and ancestral states (geographic regions, nucleotide mutations and amino acids substitutions) at internal nodes were inferred with the Augur/refine and Augur/traits modules using TreeTime (https://github.com/neherlab/treetime, version 0.11.4) [15, 28]. The “mugration” package extension of TreeTime was then used to map discrete regions to tips and infer the locations for internal nodes under a GTR model. The individual countries were used as attributes, with latitude and longitude coordinates available for national and sub-national analyses. For the inference of transmissions, note that a transmission link on the phylogeographic map can denote one or more transmission events depending on the phylogeographic inference. We first characterized SOM-BAN-1 emergence group “clades”, which we define as a group of related sequences sharing a common ancestor, with unique nucleotide mutations and translated amino acid substitutions that are shared. To further scrutinize the introduction and spread of cVDPVs within the Horn of Africa countries, we further differentiated between “chains of transmission” and “singleton introductions” among the polioviruses. We defined chains of transmission as circulating groups of polioviruses that originated from a single introduction. All poliovirus strains that clustered together were considered to belong in the same chain of transmission if their common ancestral nodes were inferred to be in a single location, whereas each individual chain of transmission was considered to be the result of a single introduction into a Horn of Africa location. Finally, the Augur/export module exports a single compiled results file required for data visualization in Auspice [15, 16]. Using the resultant annotated tree topology, the number of transitions between and within the Horn of Africa countries were counted and plotted using ggplot2 v4.0.3 [30].

The “mugration” package extension of TreeTime was also used to map discrete sample sources to tips and infer the sources for internal nodes under a GTR model. The individual source attributes were inferred based on the sample source (AFP surveillance, environmental surveillance, or contact sampling). Temporal variation in the relative frequency of poliovirus sequences by surveillance source was assessed over the study period. Sequences were categorized according to their reported source of detection, including environmental surveillance, AFP surveillance, and contact sampling. For each time interval, the proportion of sequences attributed to each surveillance source was calculated relative to the total number of sequences available, with resulting percentages visualized using a stacked area plot to illustrate changes in the relative contribution of each source over time.

We then estimated the population dynamics of cVDP2 using SOM-BAN-1 emergence group polioviruses using Delphy by measuring the relative genetic diversity (effective virus population size over time) using coalescent smoothing of the effective population size [17]. Delphy is a scalable near-real-time Bayesian phylogenetics tool for outbreaks [17].

## Results

### Spatiotemporal distribution of cVDPV2 SOM-BAN-1 emergence group polioviruses

SOM-BAN-1 emergence group was first detected in Somalia in 2017 and has been reported in 3 other countries since then (**Figure S1B, Figure 1A** and **1B**), quickly spreading from Somalia to the other countries following emergence. The temporal distribution of sequences revealed a marked shift in the relative contribution of three Horn of Africa countries between 2018 and 2025. Somalia accounted for the largest proportion of sequences during 2018–2023, consistently contributing approximately 60–100% during much of this period, although a decline was observed in 2019 and early 2021. Kenya’s contribution increased substantially from 2023, reaching approximately 60–70% during parts of 2023–2024, before declining towards the end of 2024. In contrast, Ethiopia’s contribution increased progressively from 2019, with a pronounced rise during the second half of 2024, becoming the predominant source of sequences from late 2024 through 2025. By 2025, Ethiopia accounted for nearly all sequences represented in the cVDPV2 dataset, while contributions from Somalia and Kenya had declined to minimal levels (**Figure 1C**). Overall, these findings demonstrate a temporal and geographical shift in the composition of the sequence dataset, from predominantly Somalia sequences in the earlier years of the outbreaks to increasing Kenyan representation in 2023–2024 and Ethiopian predominance from late 2024 onwards.

**Figure 1.**
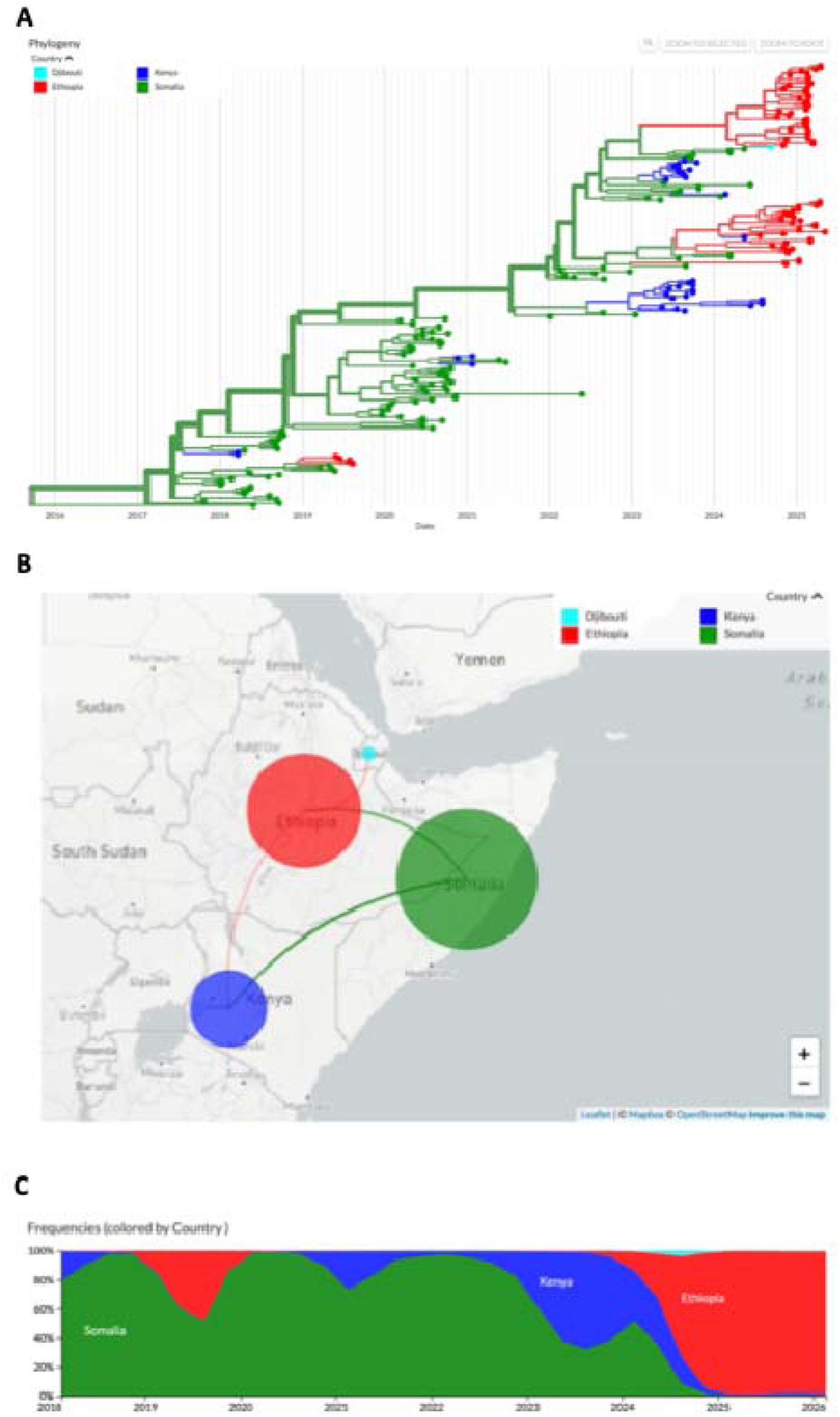
Phylogenetic reconstruction of cVDPV2 outbreaks associated with SOM-BAN-1 emergence group. (**A**) Time-resolved maximum likelihood phylogenetic tree of 234 full-length VP1 gene sequences of cVDPV2 viruses collected from four WHO AFRO and WHO EMRO countries in the Horn of Africa, with tree branches and tips color-coded as shown in the legend. (**B**) Phylogeographic reconstruction of the spread of cVDPV2 outbreak strains in the Horn of Africa showing discrete phylogeographic transmission events. Colored curved lines denote geographic transmission lines from color-coded origin to color-coded destination for exports and vice-versa in representative states (note that a transmission link can denote one or more transmission events depending on the phylogeographic inference). Circle size denotes phylogenetically inferred cVDPV2 outbreak associated with SOM-BAN-1, for example, the larger the inferred size the larger the inferred outbreak. (**C**) Temporal trends of cVDPV2 SOM-BAN-1 emergence group in four WHO countries over the study period 2017 and 2025. cVDPV2 – circulating vaccine-derived polioviruses serotype two; WHO - World Health Organization; AFRO – African Region; EMRO - Eastern Mediterranean Region.

### Characterization of the genetic diversity of cVDPV2 associated with poliovirus outbreaks in AFRO and predominance of SOM-BAN-1 emergence group in the Horn of Africa

We utilized 234 full-length VP1 gene sequences from four countries collected between 2017 and 2025 to perform phylogeographic analyses of the most predominant and most persistent emergence group in the Horn of Africa, with the aim of characterizing the evolutionary and transmission dynamics of cVDPV2 outbreaks in WHO AFRO. Discrete phylogeographic reconstruction allowed inference of the number of poliovirus imports and exports between the four countries.

Sequences from the different countries spanned the whole genetic diversity of the phylogeny, a pattern that largely reflects multiple independent introductions over time into the Horn of Africa countries (**Figure 1A**). Seeding of cVDPV2 outbreaks in these countries occurred from multiple locations to multiple destinations, with subsequent local spread, evident as distinct phylogenetic clusters of genetically similar poliovirus strains in the phylogeny (**Figure 1A** and **1B**). SOM-BAN-1 emergence group was first detected in Somalia in 2017 and has since been reported in 4 countries based on earliest epidemiological reports from outbreak investigations; the emergence group continued to circulate within the reporting countries until October 2025 (https://afrodim.shinyapps.io/new-virus-detections/), with phylogenetic predominance of Somalia over the study period (**Figure 1A** and **Figure 1C**). Somalia strains predominated the outbreaks from emergence until 2023, followed by phylogenetic predominance of Kenyan and Ethiopian strains (**Figure 1C**), a pattern which mirrors the detections (**Figure S1B**).

A molecular clock analysis of the sequences provided strong evidence for SOM-BAN-1 emergence group evolving in a clock-like manner based on root-to-tip divergence analysis (**Figure S2A**). We estimated the earliest SOM-BAN-1 emergence to a most recent common ancestor (MRCA) in February 2017, with an estimated VP1 gene evolutionary rate of 0.00983 substitutions per site per year (**Figure S2A**), which corroborates previous estimates [30], translating to the accumulation of approximately 9 substitutions per year in the VP1 gene of the SOM-BAN-1 emergence group (0.00983 substitutions/site/year by 903 base pairs for cVDPV2 VP1 gene). The earliest phylogenetic tree trunk of the cVDPV2 outbreaks originated from Somalia, consistent with SOM-BAN-1 origins in Somalia, which then seeded outbreaks in the region from 2017 onwards (**Figure 1A** and **Figure 1C**). Subsequently, Ethiopia and Kenya appear to be source populations for seeding of cVDPV2 outbreaks from 2018 onwards, but with phylogenetic predominance of Somalia.

### Understanding the transmission dynamics of cVDPV2 outbreaks based on SOM-BAN-1 emergence group

Analysis of cVDPV2 outbreaks revealed chains of SOM-BAN-1 transmission and singleton introductions into the Horn of Africa countries over the study period. A cluster of multiple virus sequences that arise from a single introduction event can be informative about virus transmission and molecular epidemiology when combined with geographic or other information. Large chains of transmission were observed in Somalia, Ethiopia, and Kenya, respectively with the largest and most persistent chain observed in Somalia (**Figure 2A**), indicating that the success of these chains of transmission was not only attributed to localized outbreaks within individual countries, but also attributable to linked regional chains of transmission that were not geographically constrained (**Figure 2A**). We then used discrete phylogeographic reconstruction of the dated phylogeny to infer the number of cVDPV2 imports and exports in the region (**Figure 2B**, **S2B**, and **S2C**) – at national and sub-national levels of observation. We inferred 14 export events (inter-Country events) over the study period (**Figure 2B**), which represent the observed introductions (circulating either as singleton introductions or chains of transmission) - **Figure 2A**. The introductions were seeded from Somalia and Ethiopia, with majority of the introductions (n=10, 71%) from Somalia to Kenya and Ethiopia, respectively (**Figure 2B**), which was also inferred from the phylogeographic reconstruction (**Figure 1B**). This underscores the important epidemiological role played by Somalia and Ethiopia in seeding cVDPV2 outbreaks in the Horn of Africa.

**Figure 2.**
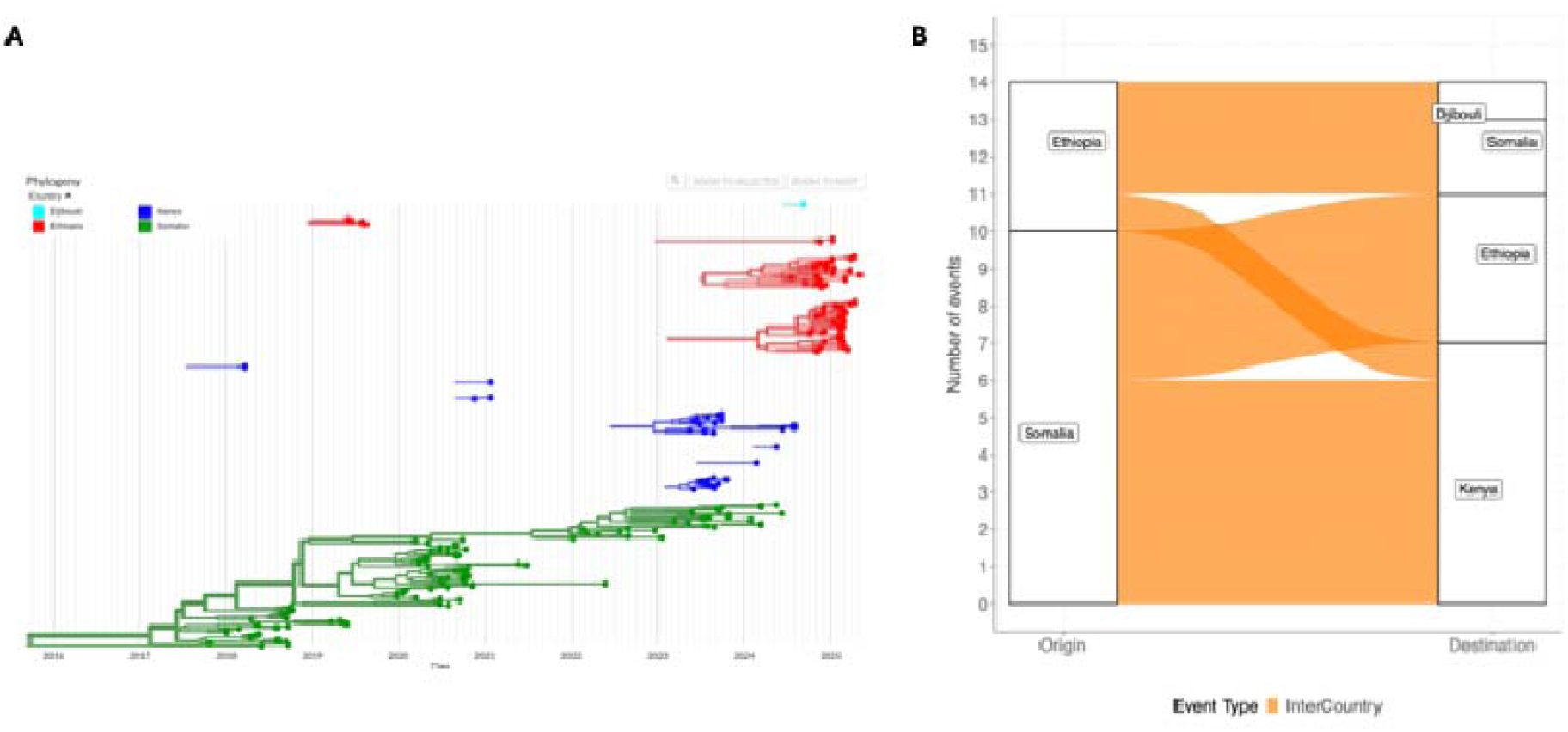
Phylogenetic reconstruction of poliovirus introductions inferred as either singleton introductions or chains of transmission of cVDPV2 SOM-BAN-1 emergence group. **(A)** Singleton introductions and chains of transmission of SOM-BAN-1 emergence group polioviruses, color-coded by country as shown in the legend. Chains of transmission are circulating groups of polioviruses originating from a single introduction. All poliovirus strains that clustered together were considered to belong to the same chain of transmission if their common ancestral nodes were inferred to be in a single location, whereas each individual chain of transmission was considered to be the result of a single introduction into a Horn of Africa country. **(B)** Sources of cVDPV2 importations and exportations in the Horn of Africa. Transition events are shown as either imports or exports by origin and destination, with events quantified as shown in the heatmap. cVDPV2 – circulating vaccine-derived polioviruses serotype two.

Majority of the chains of transmission appeared to have persisted over several epidemic seasons (**Figure 2A**), with the most persistent in Somalia supporting the concept of multi-year persistence of individual cVDPV2 chains of transmission in specific locales, contributing to continued SOM-BAN-1 circulation; persistence is also reflected in the long branches of the poliovirus phylogenies (**Figure 1A**). The persistent chains of transmission consisted of polioviruses sampled across multiple countries in the region (**Figure 2A**), which suggests that cVDPV2 persistence in the Horn of Africa region was not constrained geographically. Deeper insights into the sub-national levels of cVDPV2 outbreaks can also be elucidated from the sub-national provincial-(**Figure S2B**) and district (**Figure S2C**) transmission dynamics, which can guide targeted responses based on the sub-national dynamics, for instance, identification of important source locations for poliovirus spread and important viral transmission destinations for prioritized SIA vaccination campaigns (**Figure S3A**). For example, 68 export events were inferred at the provincial sub-national scale, with majority of the export events inferred from Somalia (Mogadishu (n=34, 50%) and Banaadir Region (n=10, 14.7%)) and Ethiopia (Oromia Region (n=9, 13%)), respectively (**Figure S2B**). At the District sub-national level, these export events were represented by transmissions from Somalia (Waberi, Mogadishu (n=42, 38.5%)) and Ethiopia (East Hararghe, Oromia Region (n=9, 8.3%)) (**Figure S2C**). Conversely, majority of import events were inferred in Banaadir (n=15, 22%) and Sool Regions (n=5, 7.4%)) of Somalia, respectively and represented by imports into Hamarweyne, Banaadir Region (n=9, 8.3%) and Urobo, Sool Region (n=6, 5.5%) at the District level of inferrence, respectively (**Figure S2B** and **S2C**).

### Transmission dynamics of polioviruses elucidated from environmental surveillance

Sequences from different sources spanned the whole genetic diversity of the phylogeny, with phylogenetic predominance of environmental surveillance sequences (**Figure 3A** and **Figure 3B**). Seeding of cVDPV2 outbreaks in the Horn of Africa countries occurred from multiple sample sources, predominatly driven by polioviruses detected and characterized from environmental surveillance.

**Figure 3.**
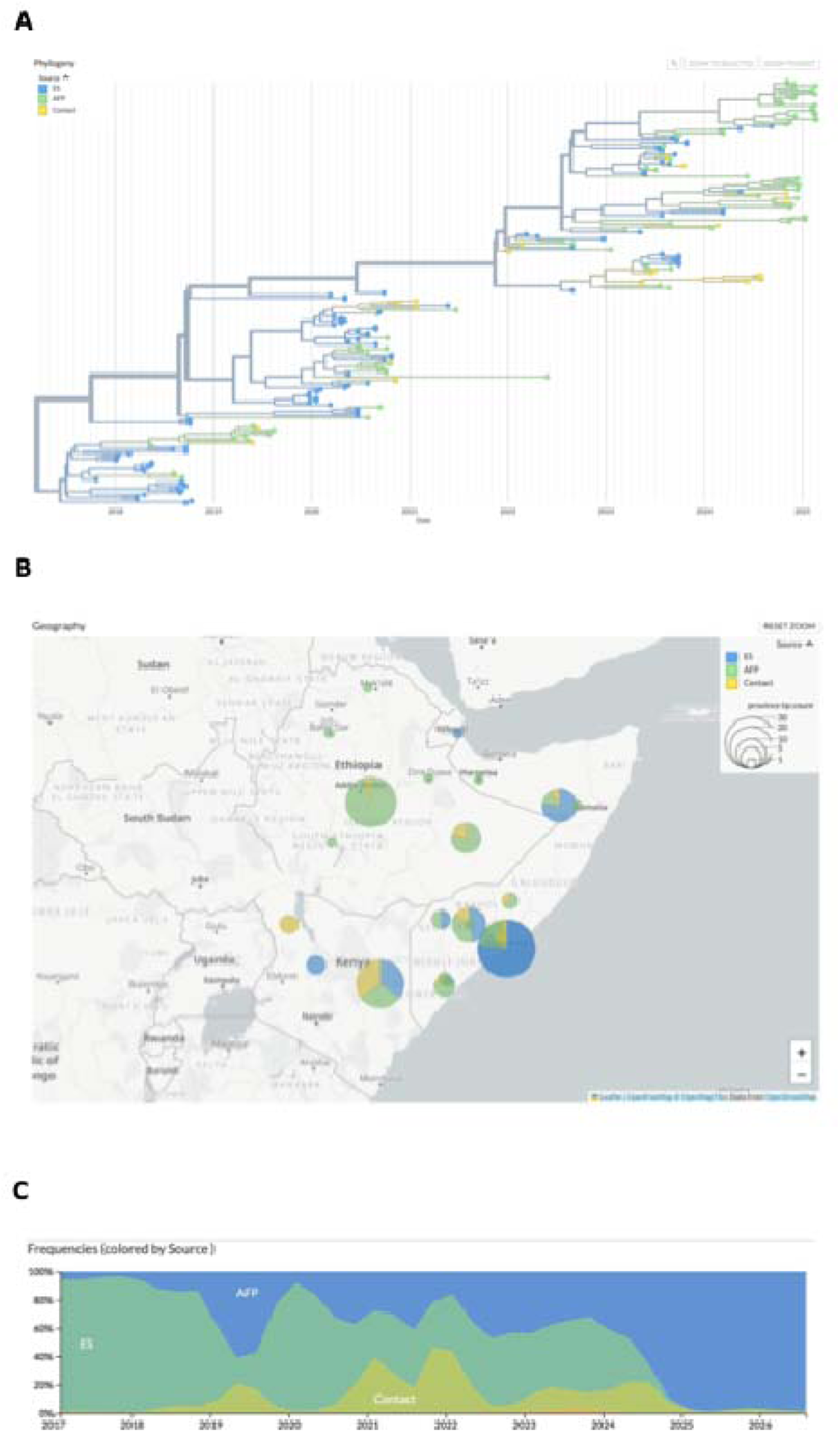
Phylogenetic reconstruction of cVDPV2 outbreaks associated with SOM-BAN-1 emergence group inferred by source of sampling. (**A**) Time-resolved maximum likelihood phylogenetic tree of 234 full-length VP1 gene sequences of cVDPV2 viruses collected from four WHO AFRO and WHO EMRO countries in the Horn of Africa, with tree branches and tips color-coded by sampling source as shown in the legend. (**B**) Phylogeographic reconstruction of the spread of cVDPV2 outbreak strains in the Horn of Africa region showing discrete phylogeographic transmission events inferred by sampling source. Colored curved lines denote transmission lines from color-coded origin to color-coded destination for exports and vice-versa in representative states (note that a transmission link can denote one or more transmission events depending on the phylogeographic inference). Circle size denotes phylogenetically inferred cVDPV2 outbreak associated with SOM-BAN-1 sampling source, for example, the larger the inferred size the larger the inferred outbreak associated with a sampling source. (**C**) Temporal trends of cVDPV2 SOM-BAN-1 emergence group in four WHO countries over the study period 2017 and 2025 inferred by sampling source. cVDPV2 – circulating vaccine-derived polioviruses serotype two; WHO - World Health Organization; AFRO – African Region; EMRO - Eastern Mediterranean Region.

The distribution of poliovirus sequences by surveillance source demonstrated substantial temporal variation over the study period (**Figure S3C**). Environmental surveillance was the predominant source of polioviruses from 2017 through 2024, generally accounting for approximately 50–95% of sequences during this period. However, its contribution fluctuated considerably, with notable declines in 2019, 2021–2022, and towards the end of 2024. AFP surveillance showed a progressive increase in its relative contribution, particularly during 2019, when it accounted for approximately 60% of sequences, subsequently becaming increasingly dominant from 2022 onwards. From late 2024, AFP surveillance became the predominant source of polioviruses, accounting for nearly 100% of sequences from 2025. Contact sampling contributed variably throughout the study period, with notable increases during 2021 and early 2022, when it represented approximately 35–45% of sequences, and smaller increases during 2019 and 2023–2024. Its contribution declined substantially after 2024 and became negligible from 2025 onwards. Overall, the findings demonstrate a transition in the relative composition of cVDPV2 sequences from predominantly environmental surveillance during the earlier years to predominantly AFP surveillance from 2025 onwards, while contact sampling made intermittent contributions.

A total of 53 exportation events were inferred at the regional levels, with the majority of the exportations inferred from environmetal surveillance (n=30, 56.6%) and AFP (n=16, 30.2%) (**Figure S2B**). Conversely, majority of importation events were inferred from AFP (n=28, 52.8%) and contact sampling (n=15, 28.3%), respectively.

### Elucidating the population dynamics of cVDP2 outbreaks using SOM-BAN-1 emergence group

Coalescent reconstruction of the SOM-BAN-1 cVDPV2 emergence group in the Horn of Africa during the study period using the Delphy Skygrid model revealed: (i) multi-year cyclical fluctuations in relative genetic diversity between 2017 and 2025 following an initial rapid expansion during its 2017 emergence in Banadir, Somalia; (ii) three distinct peaks in late 2017, early 2021, and late 2023; and (iii) intervening genetic bottlenecks in mid-2019 and early 2022 associated with outbreak response campaigns, before stabilizing at a moderate plateau from mid-2024 up to the final detection of the lineage in October 2025 (**Figure 4**). Overall, posterior median values remained consistently below 10 days, reflecting persistent, cryptic circulation and periodic cross-border exportation rather than explosive epidemic expansion.

**Figure 4.**
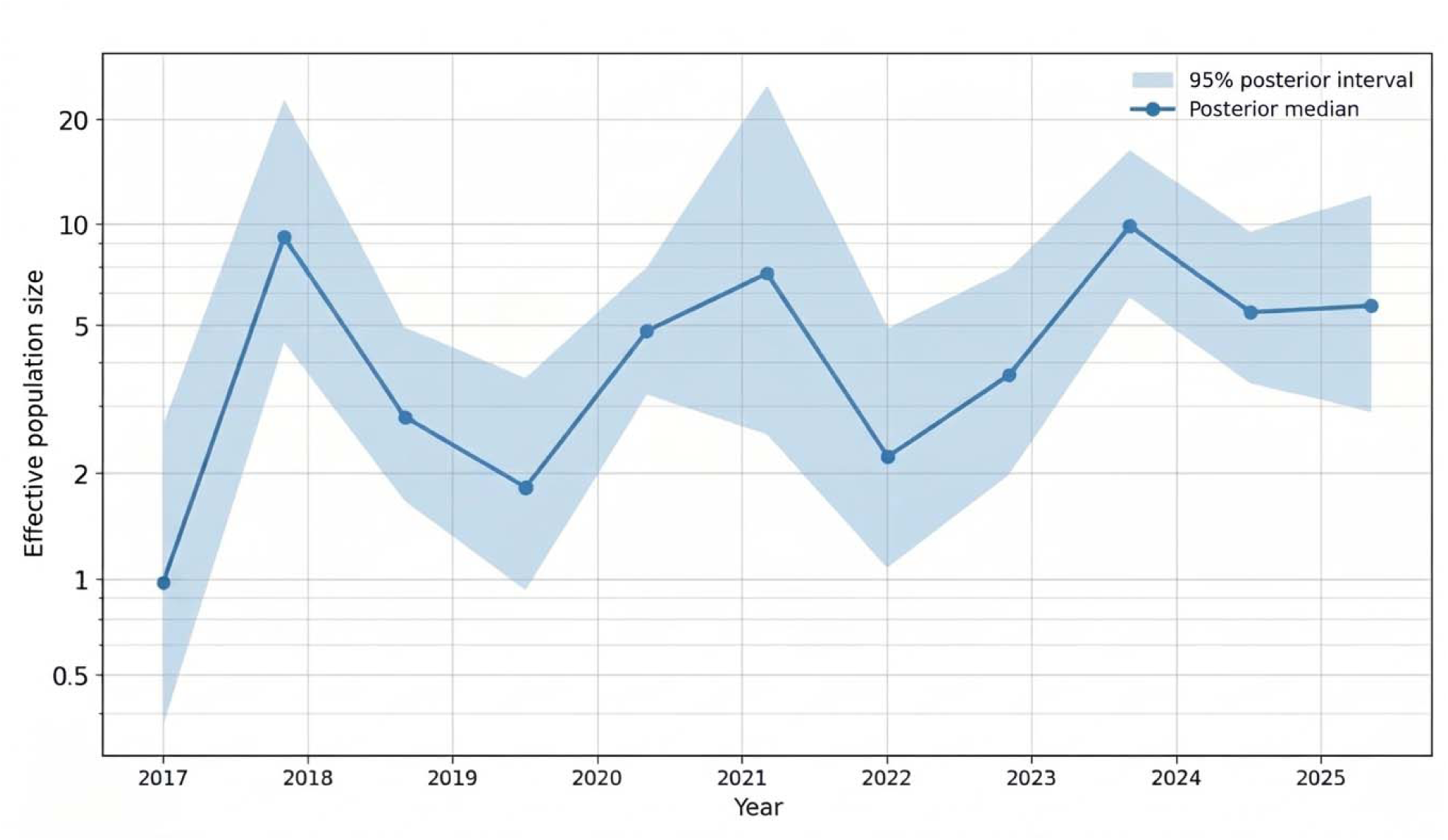
Estimates of the relative genetic diversity through time for cVDPV2 SOM-BAN-1 emergence group from the Horn of Africa region, resolved through coalescence analysis.

## Discussion

We observed that cVDPV2 strains from different countries in the Horn of Africa spanned the overall genetic diversity of the cVDPV2 phylogeny, which reflects multiple independent introductions over time into the region. Seeding of these polioviruses into the countries in the region occurred from multiple locations to multiple destinations, with subsequent local spread, evident as distinct phylogenetic clusters of genetically similar poliovirus strains in the regional phylogeny. Temporal and geographical shift in the composition of the sequence dataset, from predominantly Somali sequences in the earlier years to increasing Kenyan representation in 2023–2024 and Ethiopian predominance from late 2024 onwards was observed. Phylogeographic analyses revealed SOM-BAN-1 emergence group origins from Somalia, with subsequent spread to other regional countries and subsequent seeding of cVDPV2 outbreaks through other epidemiologically important countries, but with phylogenetic predominance of Somalia. Additionally, the regional transmission dynamics of cVDPV2 was characterized by the circulation of multiple chains of transmission in multiple neighboring countries, with poliovirus spread characterized by localized virus evolution and circulation of regionally linked chains of transmission. Coalescent reconstruction of revealed multi-year oscillations in relative genetic diversity between 2017 and 2025 following an initial rapid expansion after emergence, with three distinct peaks in late 2017, early 2021, and late 2023 and intervening genetic bottlenecks in mid-2019 and early 2022 associated with outbreak response campaigns, before stabilizing at a moderate plateau from mid-2024. Overall, posterior median values remained consistent, reflecting persistent, cryptic circulation and periodic cross-border exportation rather than explosive epidemic expansion.

SOM-BAN-1 emergence group, which has been the most predominant and persistent emergence group in the region, continued to evolve since emergence in 2017, subsequently spreading to neighboring countries in the region. Additionally, phylogenetic tree trunks of cVDPV2 outbreaks originated predominantly in Somalia, consistent with viral origins of SOM-BAN-1 emergence group in Somalia in 2017, which then seeded cVDPV2 outbreaks in the region. Furthermore, our analyses revealed chains of transmission of cVDPV2 and singleton introductions into the region, with the largest transmission chains observed in Somalia, Ethiopia, and Kenya, which indicates that the success of these chains of transmission was not only due to localized outbreaks in different countries but also due to linked chains of transmission in neighboring countries that were not geographically constrained. Persistence of chains of transmission, also reflected in long branches of the poliovirus phylogenies, supports the concept of multiyear persistence of individual cVDPV2 outbreak transmission chains in specific locales possibly reflecting potential gaps in outbreak response and surveillance. Wider and deeper sampling and analysis of sequences from additional cVDPV2 outbreaks is required for improved understanding of the evolutionary and transmission dynamics of this emergence group. Additionally, phylogeographical analyses provided deeper insights into the sub-national transmission dynamics of cVDPV2 identifying important source locations and destinations for poliovirus spread, which can be important for identiying regions for prioritization for SIA vaccination campaigns.

Investigation of the genetic relatedness of viruses is important in the characterization of the patterns of introduction, persistence and geographic spread of poliovirus. Through phylogeographic analyses, the patterns of poliovirus introduction (whether single or repeated), persistence and spread can be elucidated using phylogeographic models, for example, discrete trait analyses that provide geographic histories of viral movement and associated rates [15–17]. These approaches have been applied to WPV1 sequences from Pakistan and Afghanistan, where WPV1 persists through complex dynamics of evolution and spread [31]. Genomic analysis has also been applied to distinguish between cVDPV outbreaks arising from local transmission and those that are likely to be linked to chains of transmission [9, 14]. Here, through detailed genomic epidemiology, we extend these observations, where we observed the circulation of linked chains of transmission in multiple countries and singleton introductions into countries. Our analysis also revealed sustained persistence of multiple chains of transmission, although increased sampling is required to confirm that cVDPV2 outbreak strains from other localities are not interspersed within these chains of transmission. Prolonged transmission of a cVDPV2 (lasting ≥12 months from first to most recent detection) was observed in Somalia, Ethiopia and Kenya during January 2023-June 2024 [9], remaining as prevalent as those during previous reporting periods [32, 33]. The spread of cVDPV2 emergence groups such as SOM-BAN-1 outside the country of first detection, reveals gaps in the effectiveness and timeliness of outbreak responses and the need for novel strategies to guide implementation of more strategic and effective vaccination campaigns.

Coalescent reconstruction of the SOM-BAN-1 emergence group revealed multi-year oscillations in relative genetic diversity between 2017 and 2025, following an initial rapid expansion during its 2017 emergence in Banadir, Somalia, capturing the initial emergence and establishment of SOM-BAN-1 in Banadir Province, Somalia (first detected in October 2017 following the 2016 trivalent-to-bivalent OPV switch) and its subsequent cross-border spread into Kenya and the Somali Region of Ethiopia. This was followed by contraction in relative genetic diversity to its lowest trough in mid-2019, which aligned with intensive monovalent OPV2 (mOPV2) outbreak response campaigns that temporarily curtailed transmission, followed by a resurgence peaking in early 2021, possibly driven by COVID-19 disruptions to routine immunization and SIAs. Following a second genetic bottleneck in early 2022, SOM-BAN-1 experienced its highest median genetic diversity peak in late 2023, coinciding with sustained transmission in security-compromised, zero-dose districts of South-Central Somalia (such as Banadir and Jubbaland) and renewed cross-border detections in Kenya, Ethiopia and Djibouti (https://afrodim.shinyapps.io/new-virus-detections/). The stabilization in relative genetic diversity from mid-2024 through 2025 reflected continued low-level persistence of SOM-BAN-1 in hard-to-reach reservoirs balanced against expanded novel OPV2 (nOPV2) vaccination rounds across Somalia and neighboring Horn of Africa countries. Overall, posterior median values remained consistently below 10 days, reflecting persistent low-level regional circulation balanced against ongoing vaccination efforts prior to apparent interruption in late 2025. These insights could be useful in public health strategies for future emergence groups of cVDPV2 like the newly emerged SOM-BAY-1 (https://afrodim.shinyapps.io/new-virus-detections/) emergence group in the Horn of Africa, for example, in guiding the timings for vaccinations and SIAs.

Despite a globally coordinated switch from tOPV to bOPV in April 2016 in routine and SIAs through the global withdrawal of serotype two OPV, cVDPV outbreaks have been increasing in recent years, with new cVDPV2 outbreaks continuously reported in multiple countries [8–10]. Prolonged community circulation of the vaccine strain leads to reversion to neurovirulence, seeding new emergences, with prolonged undetected circulation of poliovirus in areas with weak surveillance systems creating gaps in surveillance, majority of infections being either asymptomatic or resulting in mild illness. Gaps in poliovirus surveillance can delay outbreak response activities, providing a longer opportunity for viruses to spread [9, 13]. Additionally, there is increased risk of the locally evolved cVDPV2 polioviruses seeding outbreaks in other regions as recently reported in EURO, where cVDPV2 isolates from environmental samples in five EURO countries were genetically linked to cVDPV2 emergence first reported in Nigeria [12].

Environmental surveillance of wastewater for poliovirus was introduced to complement AFP surveillance, enabling early detection in communities with poliovirus transmission without evidence of clinical cases. Environmental surveillance involves testing sewage runoff for the presence of poliovirus. Because the majority of cases of poliomyelitis are asymptomatic, but all infected people shed virus in their stool, environmental surveillance has the benefit of allowing us to detect the presence of polio in an area before any case of paralysis appears [19, 21, 34]. Here, phylogeographic analysis of sequence data generated from environmental surveillance, AFP, and contact samples revealed phylogenetic predominance of environmental surveillance sequences and seeding of cVDPV2 outbreaks in the Horn of Africa countries predominatly driven by viruses detected and characterized from environmental surveillance. Therefore, the strengthening of environmental surveillance is important to detect silent poliovirus circulation in communities by testing sewage and wastewater before any cases of paralysis occur [21, 34]. This will important for early outbreak detection, higher sensitivity for detection of polioviruses, tracking transmission pathways, evaluating vaccination campaigns, and certifying of polio-free status [19, 21, 34].

Efforts are underway to strengthen surveillance systems and improve the capacity to confirm cVDPV outbreaks by increasing the number of laboratories accredited by Global Poliovirus Laboratory Network (GPLN) to perform genomic sequencing [6]. Analysis of genetic relatedness of poliovirus sequences can act as a good marker of the progress towards eradication, filling such gaps in surveillance. For example, reduction in the number of cVDPV2 outbreaks may be indicative of impact of vaccination campaign, monitored through the reduction of chains of transmission [10] as observed for SOM-BAN-1. We observed the circulation of multiple chains of transmission during the study period, associated with the circulation of cVDPV2 chains of transmission of the SOM-BAN-1 emergence group. Monitoring of the number of circulating chains of transmission over specific time periods and the inferred branch lengths of cVDPV2 outbreak phylogenies at regional, countrywide, and sub-national levels may be indicative of impact of vaccination campaign.

The current GPEI target was to stop all cVDPV transmission by the end of 2026. However, the continued circulation of cVDPVs highlights the need for 1) increased urgency to implement prompt, high-quality SIAs upon detection of new cVDPV outbreaks and 2) enhanced efforts, such as more engagement with humanitarian nongovernmental organizations, to vaccinate children in security-compromised areas and in hard-to-reach communities. Regular updates of the poliovirus surveillance sequence data using Nextstrain and Delphy visualization tools, incorporating phylogeographic analyses output as presented in our work, may provide insights into changes in poliovirus epidemiology, which can in turn guide the implementation of more strategic and effective SIAs. These analyses may be particularly relevant to halt the ongoing transmission of cVDPVs in the AFRO, seeding of cVDPV2 outbreaks from AFRO to other regions, and control of importation of WPV1 into AFRO, through improved surveillance and timely and effective outbreak response, achieving the GPEIs global poliovirus eradication efforts.

## Data Availability

All data produced utilised in this study are available online at https://extranet.who.int/polis/Account/Login.

## Supplementary Figures

**Figure S1.**
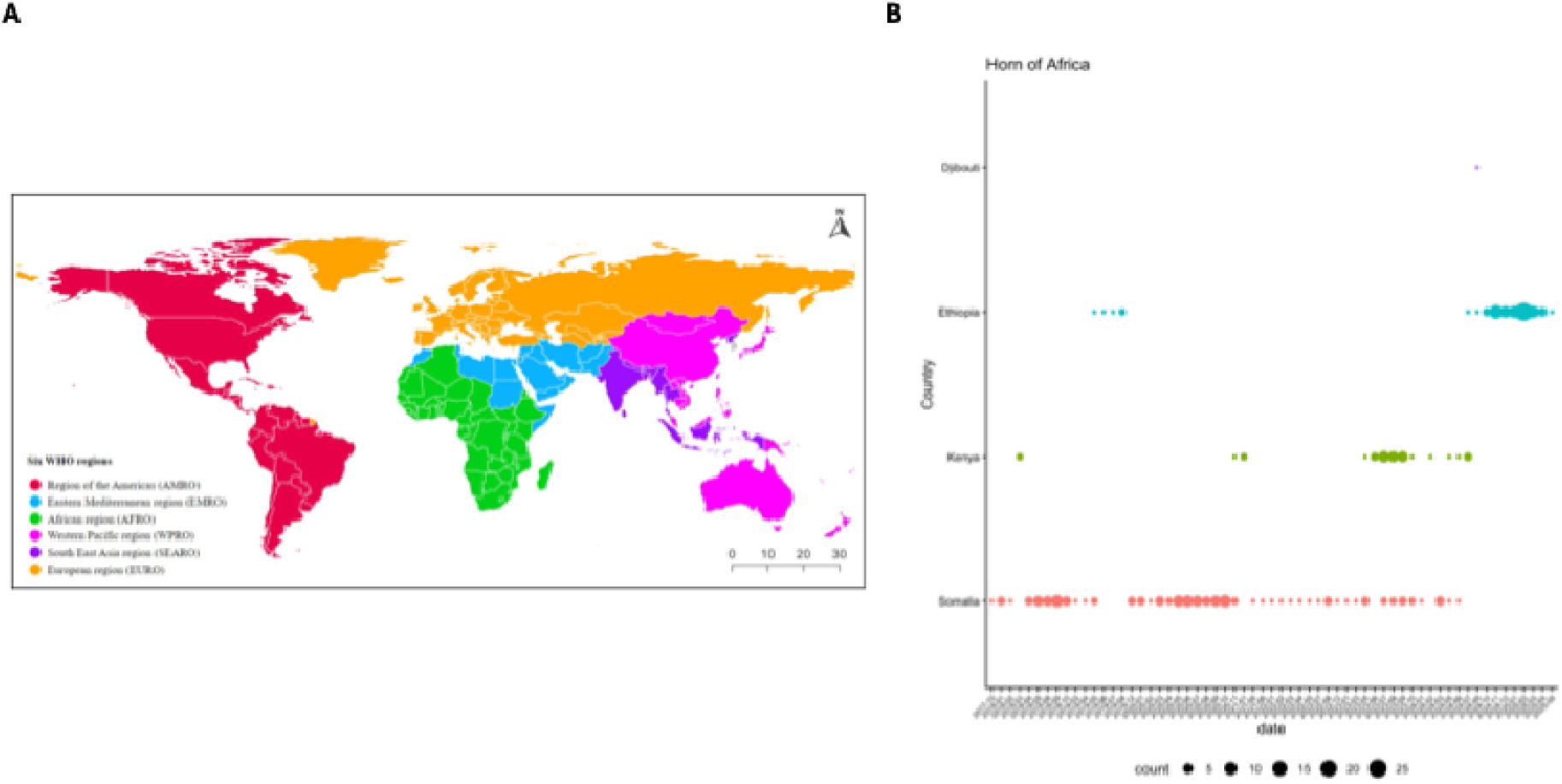
World Health Organization (WHO) global regions and spatiotemporal distribution of cVDPV2 sequences in the Horn of Africa of the African Region (AFRO). **(A)** Map of WHO global regions, 2026. Country assignments to WHO regions are based on administrative designation rather than strictly geographic criteria. **(B)** Temporal distribution of cVDPV2 SOM-BAN-1 emergence group sequences collected from WHO AFRO countries in the Horn of Africa between October 2017 and May 2025. The number of sequences by country are shown between 2017 and 2025 as represented in the legend. cVDPV2 – circulating vaccine-derived polioviruses serotype two. WHO - World Health Organization; AFRO – African Region.

**Figure S2.**
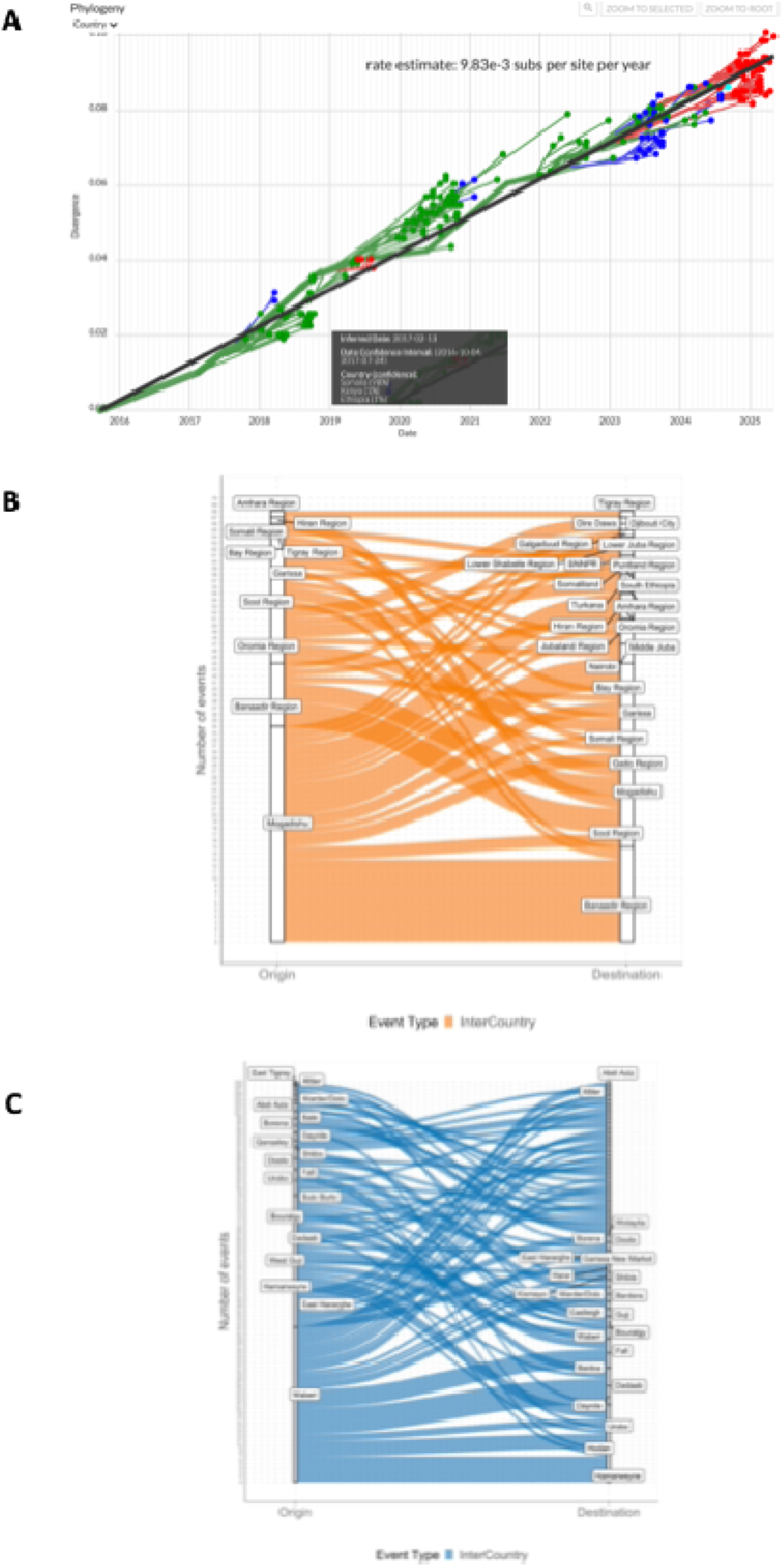
Evolutionary and transmission dynamics of SOM-BAN-1 poliovirus. **(A)** Root-to-tip divergence plot for cVDPV2 SOM-BAN-1 emergence group polioviruses collected from WHO AFRO and EMRO countries. (**B**) Transition events (imports and exports) between WHO countries in the Horn of Africa shown as “inter-Country” events between countries inferred at the provincial level of observation. (**C**) Transition events (imports and exports) between WHO countries in the Horn of Africa shown as “inter-Country” events between countries inferred at the district level of observation. cVDPV2 – circulating vaccine-derived polioviruses serotype two. WHO - World Health Organization; AFRO – African Region.

**Figure S3.**
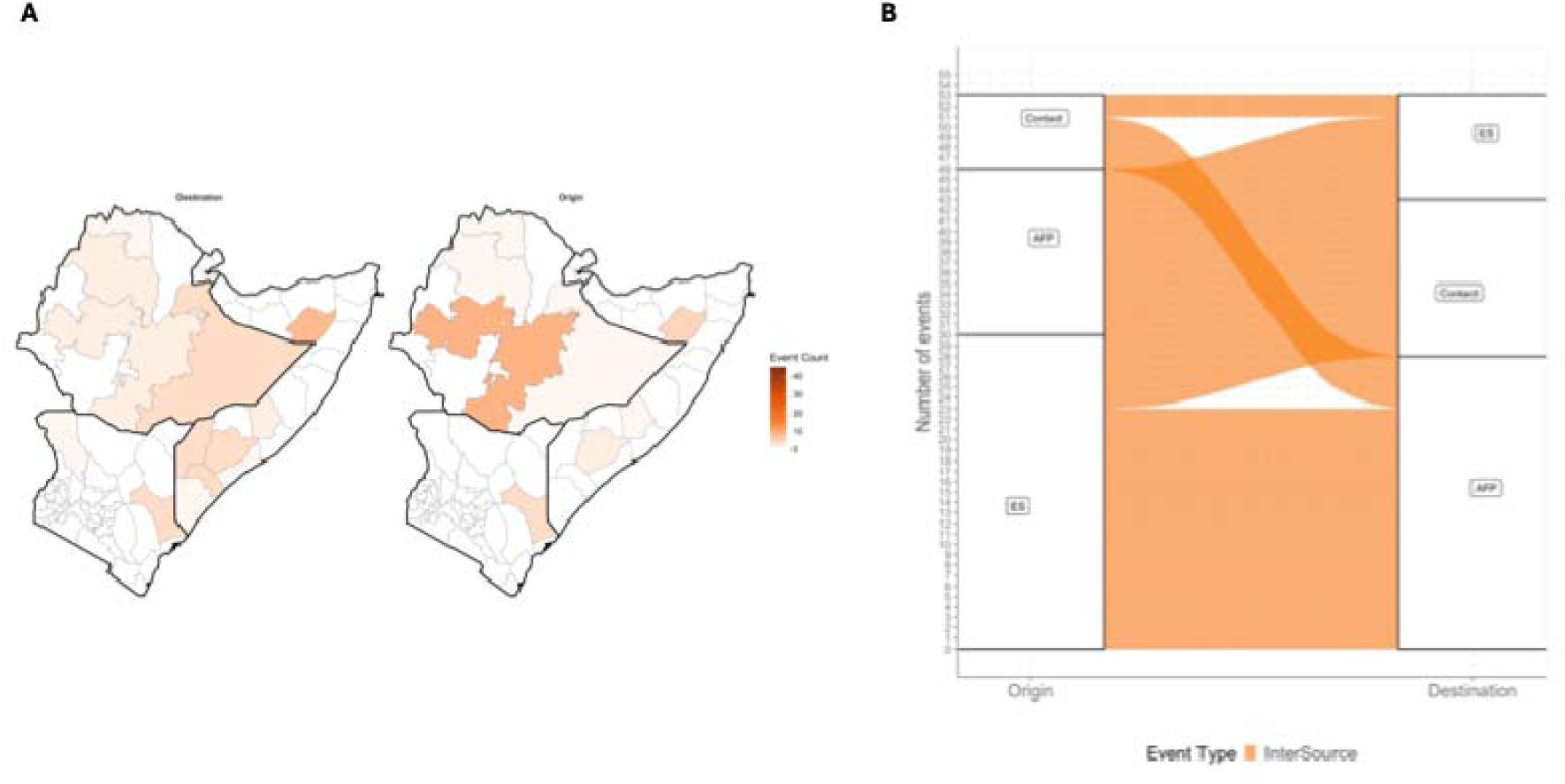
**(A)** Origins and destinations of cVDPV2 importations and exportations in the Horn of Africa inferred from phylogeographic reconstruction and visualized on the Horn of Africa regional map. **(B)** Sources of cVDPV2 importations and exportations in the Horn of Africa inferred from poliovirus sampling source. Transition events are shown as either imports or exports by origin and destination, with events quantified as shown in the Y-axis. cVDPV2 – circulating vaccine-derived polioviruses serotype two.

